# Insulin resistance potentiates the effect of remnant cholesterol on cardiovascular mortality in individuals without diabetes

**DOI:** 10.1101/2023.04.14.23288605

**Authors:** Arsenio Vargas-Vázquez, Carlos A. Fermín-Martínez, Neftali Eduardo Antonio-Villa, Luisa Fernández-Chirino, Daniel Ramírez-García, Gael Dávila-López, Juan Pablo Díaz-Sánchez, Carlos A. Aguilar-Salinas, Jacqueline A. Seiglie, Omar Yaxmehen Bello-Chavolla

## Abstract

**BACKGROUND:** Remnant cholesterol (RC) and insulin resistance (IR) have been independently associated with cardiovascular risk.

**OBJECTIVES:** We evaluated the mediating role of IR in the risk conferred by RC on atherosclerotic cardiovascular disease (ASCVD) mortality.

**METHODS:** Analysis of the National Health and Nutrition Examination Survey (NHANES-III/IV), including 16,201 individuals ≥20 years without diabetes and with complete mortality data. RC levels were calculated using total cholesterol, non-HDL-c, and LDL-c estimated with Sampson’s formula; IR was defined as HOMA2-IR≥2.5 and ASCVD mortality as a composite of cardiovascular and cerebrovascular mortality. Multiple linear regression was used to assess the relationship between HOMA2-IR and RC and competing-risk models to assess their joint role in ASCVD mortality. Causally ordered mediation models were used to explore the mediating role of IR in RC-associated ASCVD mortality.

**RESULTS:** We identified an association between higher HOMA2-IR and higher RC levels. The effect of IR on ASCVD mortality was predominant (sHR 1.68, 95%CI 1.42-1.98) and decreased at older ages (sHR 0.993, 95%CI 0.991-0.995) compared to RC (sHR 0.965, 95%CI 0.826-1.126). Higher risk of ASCVD mortality was observed in individuals with IR but normal RC (sHR 1.19, 95%CI 1.01-1.41) and subjects with IR and high RC (sHR 1.27, 95%CI 1.07-1.50), but not in subjects without IR but high RC. In mediation models, HOMA2-IR accounted for 86.5% (95%CI 78.8-100.0) of the effect of RC levels on ASCVD mortality.

**CONCLUSIONS:** Our findings suggest that RC potentiates the risk of ASCVD mortality through its effect on whole-body insulin sensitivity, particularly among younger individuals.

**CONDENSED ABSTRACT:** This study aimed to assess the role of insulin resistance (IR) in the association between remnant cholesterol (RC) and atherosclerotic cardiovascular disease (ASCVD) mortality. Using data from NHANES-III/IV, the study found that the effect of IR on ASCVD mortality was predominant compared to RC. Individuals with IR but normal RC or IR with elevated RC had higher risk of ASCVD mortality compared to subjects with elevated RC without IR. Overall, IR accounted for 86.5% of the effect of RC on ASCVD mortality. RC increases the risk of ASCVD mortality through its effect on insulin sensitivity, particularly among younger individuals.

**PERSPECTIVES:** *COMPETENCY IN PATIENT CARE AND PROCEDURAL SKILLS:* In patients with elevated remnant cholesterol, insulin resistance potentiates its effect on atherosclerotic cardiovascular disease mortality.

*TRANSLATIONAL OUTLOOK:* Additional research from randomized clinical trials is needed to evaluate whether concomitant therapies to improve insulin sensitivity in patients with residual cardiovascular risk may provide an additional benefit in reducing cardiovascular disease burden and mortality.

**GRAPHICAL ABSTRACT – CENTRAL IMAGE:** 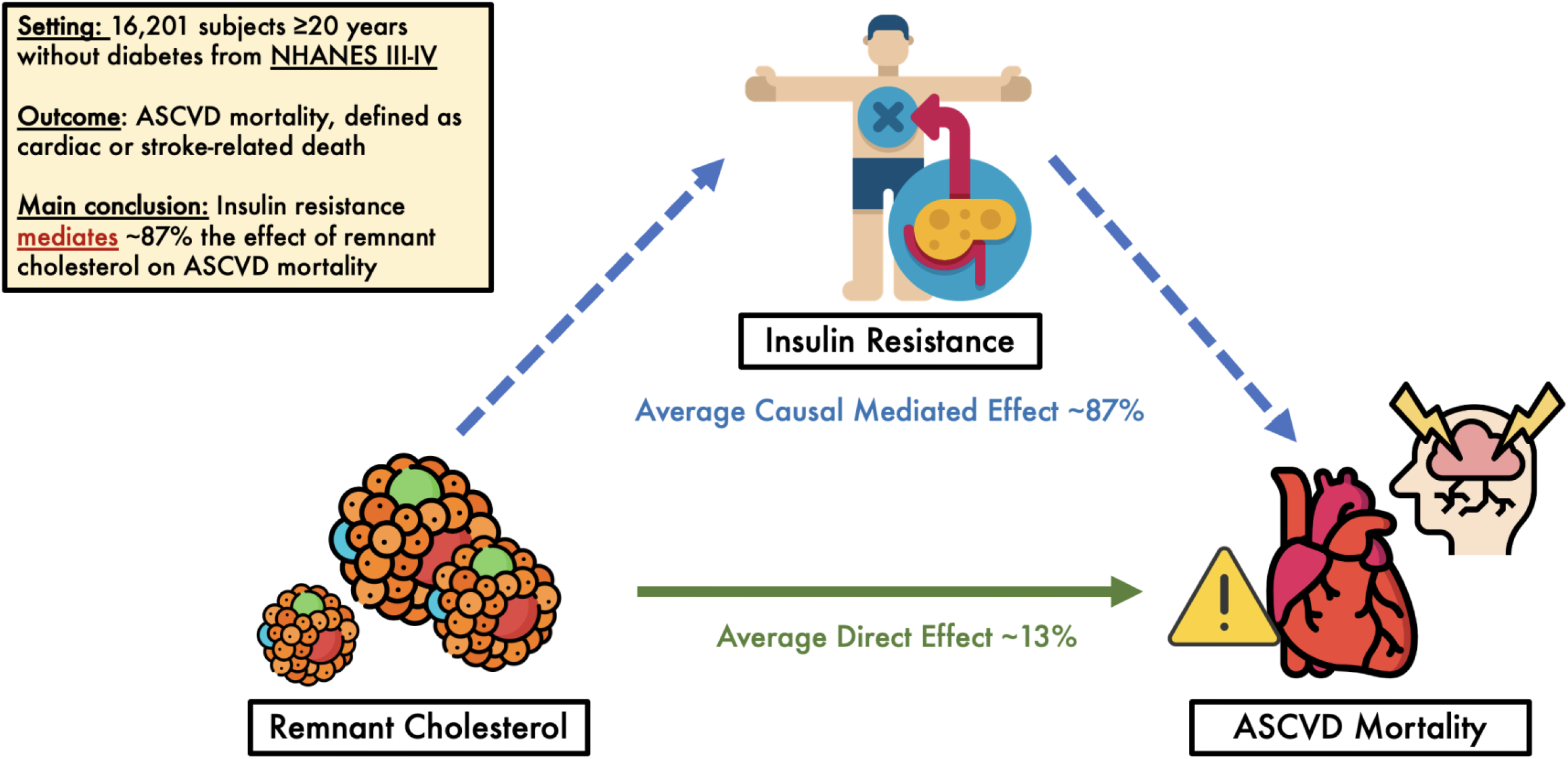

## INTRODUCTION

Triglyceride-rich lipoprotein (TRL) remnants, also known as remnant cholesterol (RC), are the products of triglyceride-rich lipoprotein particles that have been metabolized primarily by lipases to produce cholesterol-enriched particles^1,2^. During a state of whole-body insulin resistance (IR), dyslipidemia may occur with concurrent hypertriglyceridemia, decreased HDL levels, and changes in lipoprotein composition^3,4^. Hyperinsulinemia and central obesity, which typically accompany IR, condition overproduction of VLDL^4,5^. Recent studies have shown a causal association between triglycerides, TRL, and RC, with an increased risk of cardiovascular disease^6,7^. Furthermore, increased RC levels may contribute significantly to residual cardiovascular risk in patients receiving optimized low-density lipoprotein (LDL-C) lowering therapy^8^. Cholesterol remnants may be involved in the development of atherosclerosis and ischemic heart disease through accumulation of cholesterol in the arterial wall, endothelial dysfunction, impaired vasodilation, proinflammatory response, and plaque rupture^3,6,9,10^. Despite this, the complexity of TRL and RC metabolism and its measurement has yielded inconsistent results on the effect of its reduction in cardiovascular outcomes in randomized controlled trials, indicating a need to further understand additional pathophysiological mechanisms involved in this process^3^.

Given the role of IR in increasing RC and the known impact of IR in increasing risk for cardiovascular disease, it is of interest to explore whether IR participates in the causal pathway that links RC with increased cardiovascular risk and mortality^11,12^. To this end, it is useful to implement approaches that allow us to deconstruct the total effect of RC on cardiovascular mortality, in a direct effect explained by the atherosclerotic mechanisms attributable to RC and an indirect effect attributable to the effect of RC on IR, which subsequently increases ASCVD mortality^13,14^. In this study, we aim to estimate the indirect effect attributable to IR of the total effect of RC on cardiovascular mortality in subjects without diabetes and to identify the main predictors of RC concentration to better understand how insulin resistance modulates cardiovascular risk associated with RC independent of other atherogenic lipoprotein profiles.

## METHODS

### Study population and design

We used data from the National Health and Nutrition Examination Survey (NHANES), a survey program designed to assess the health and nutritional status of adults and children in the United States. Specifically, we used data from NHANES III, a national probabilistic sample of 39,695 individuals carried out between 1988 and 1994 in two phases and NHANES-IV, which was conducted between 1999 and 2018. Given that the estimation of IR using HOMA2-IR in individuals with diabetes may be influenced by insulin treatment and glycemic control^15^, we focused our analyses in subjects ≥20 years without diabetes, defined as a previous diagnosis of diabetes and/or glycosylated hemoglobin ≥ 6.5%, and/or fasting glucose ≥ 126mg/dL. We included participants with complete demographic, anthropometric and biochemical data as well as linked mortality follow-up in whom RC estimation was feasible. A detailed overview of included subjects is presented in **Figure 1**. All proceedings related to this work were registered and approved by the Research Committee at Instituto Nacional de Geriatría, project number DI-PI-006/2020.

**Figure 1.**
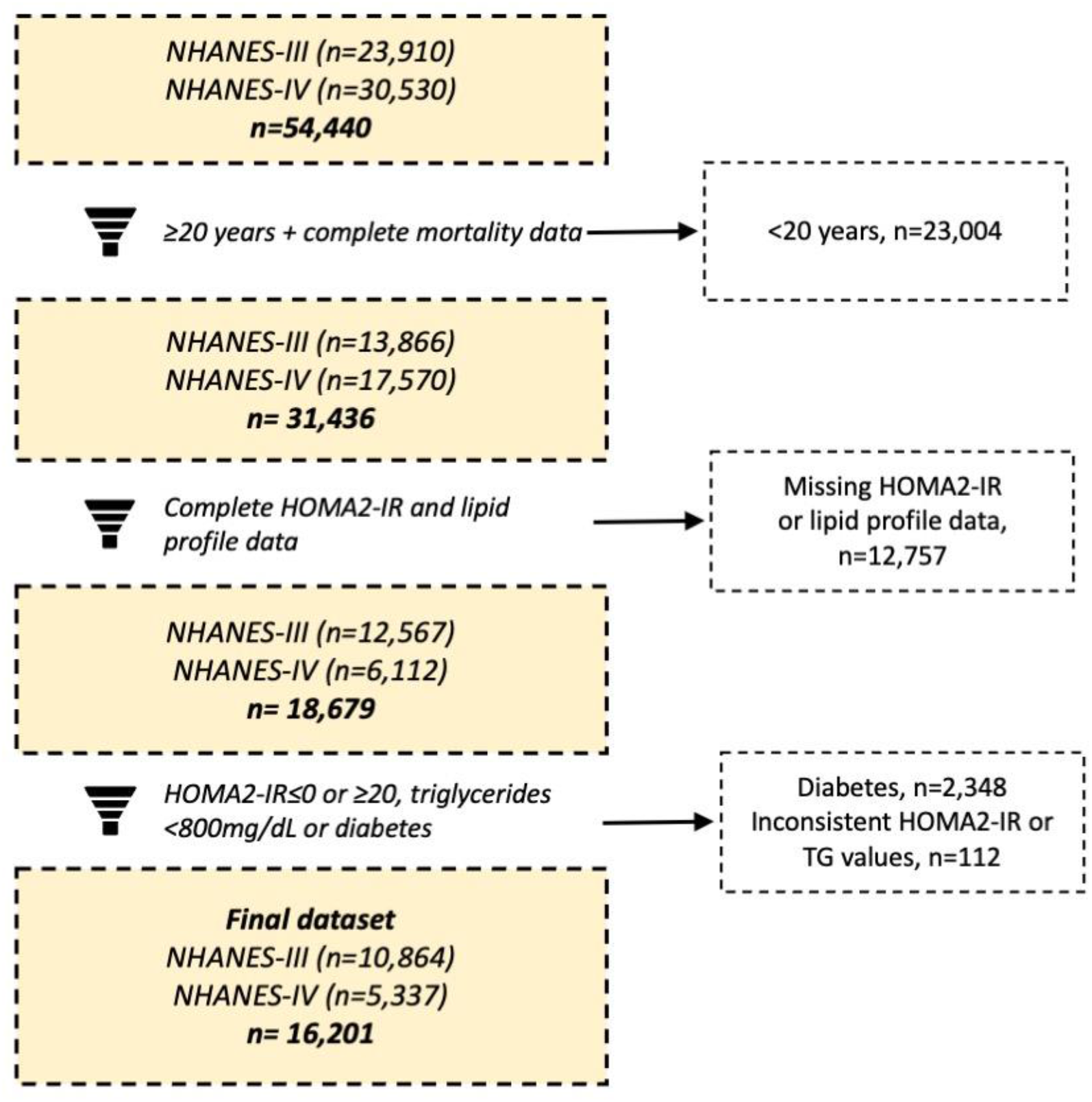
Flowchart of study participant selection derived from the original sample in NHANES-III and NHANES-IV

### Variables and definitions

#### Assessment of insulin resistance

We used the Homeostatic Model Assessment to derive indices of IR (HOMA2-IR), beta cell function (HOMA2-B), and insulin sensitivity (HOMA2-S). The HOMA2 model was estimated using fasting glucose and C-peptide levels with the calculator available at https://www.dtu.ox.ac.uk/homacalculator/. HOMA2-IR values ≥2.5 were used to define IR^16,17^. We eliminated subjects with HOMA2-IR values ≥20.0 (>99th percentile), since these values may indicate underlying disease associated with extreme IR (e.g., lipodystrophies).

#### Remnant cholesterol estimation

LDL cholesterol (LDL-c) was estimated using two methods. Sampson’s formula (LDL_S_) was developed as an estimate of LDL-C in patients with hypertriglyceridemia or low triglyceride levels^18,19^, as described below:

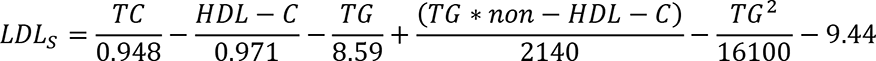

As a sensitivity analysis, we also utilized Martin’s formula (LDL_M_) which uses an adjustable factor of the TG:VLDL-C ratio (*x*_*i*_), specified according to TG levels, total cholesterol (TC) and HDL-C. As described in the following formula:

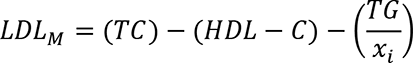

Given that the estimation of RC is dependent on LDL-C estimation, we used both LDL_S_ and LDL_M_ to estimate RC using the following formula:

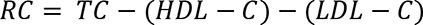

Considering that formulas to calculate LDL-C, such as Martin’s and Sampson’s show a low performance in severe hypertriglyceridemia, we decided to eliminate subjects with TG ≥800mg/dL from the analysis ^19,20^. We used RC values ≥30mg/dL to define elevated RC^21^. All analyses herein presented are evaluated using LDL-C estimated with Sampson’s formula, whilst estimates with Martin’s formula are available in **Supplementary Material**.

#### Outcome variables

1. All-cause mortality, and cause-specific mortality (heart disease, cerebrovascular disease, malignancies, accidents, diabetes, influenza or pneumonia, Alzheimer’s, chronic obstructive pulmonary disease, other causes) were obtained using national death registry linkage with NHANES data. Follow-up time was defined as time in months from interview date to confirmation of mortality status in 2018.
2. Atherosclerotic cardiovascular disease (ASCVD) mortality was defined as a composite of mortality associated with either heart and/or cerebrovascular disease and was defined as the main outcome measure of our study. To account for the specific risk attributable to this cause of death, all mortality analyses were fitted considering competing risks.

#### Additional covariates

We included variables pertaining to personal medical history (hypertension, heart failure, ischemic heart disease, cerebrovascular disease), gender, age, ethnicity (African American, Mexican American, Caucasian, and others). Detailed procedures for biochemical and anthropometric measurements have been previously described^22,23^. The following variables were retrieved for our analysis: glycated hemoglobin (HbA1c), total cholesterol, HDL cholesterol (HDL-C), triglycerides, weight, height, and waist circumference. Body mass index (BMI) was calculated as weight in kilograms divided by the square of height in meters. Prediabetes was defined according to ADA definitions as HbA1c ≥5.7%.

### Statistical analyses

#### Relationship between insulin resistance and remnant cholesterol

To study the relationship between RC and IR we fitted a linear regression model using HOMA2-IR as a dependent variable, adjusted for age, gender, ethnicity, BMI, smoking, LDL and HDL cholesterol, and number of comorbidities. Model assumptions were validated using graphical methods and specific statistical tests for model residuals; model selection was conducted by minimization of the Bayesian Information Criterion (BIC). To account for the possibility of a bidirectional relationship, we also explored how RC levels varied according to levels of IR using linear regression, further stratifying subjects according to LDL-c levels and according to TG levels (≥200mg/dL) for all models.

#### Mortality rates according to remnant cholesterol and insulin resistance levels

We calculated mortality rates by standardizing rates to deaths per 1,000 person-years and their corresponding 95% confidence intervals. These rates were evaluated for all-cause, heart, cerebrovascular and ASCVD mortality. Rates were subsequently stratified based on IR (HOMA2-IR ≥2.5), elevated RC (≥30mg/dL) and the combination of both criteria as exploratory analyses; however, for adjusted mortality analyses all variables were treated as continuous variables.

#### Association of remnant cholesterol and insulin resistance with cardiovascular mortality

To evaluate the risk of ASCVD mortality associated with both RC and IR we used a Fine & Gray competing risk regression model, calculating follow-up time date from the date of initial evaluation until cardiovascular death, death by any other cause or censorship, whichever occurred first. First, we fitted a model with only lipoprotein levels (LDL-C, HDL-C, RC), which were subsequently adjusted by age and sex and then a fully adjusted model with additional adjustments for BMI, HbA1c, ethnicity, smoking, hypertension, and number of comorbidities. We then fitted a model that incorporated HOMA2-IR into the model to explore its modifying role in the association of lipoproteins with ASCVD mortality. To further characterize the increasing risk of cardiovascular mortality with increasing values of RC and HOMA2-IR we created an indicator variable with HOMA2-IR values and RC and created four categories: Subjects without IR and normal RC, IR with normal RC, No IR and elevated RC and IR with elevated RC and explored fully adjusted risk for cardiovascular disease for these categories using competing risk analyses. We also used the *simPH* R package to simulate the sub-distribution hazard ratio for increasing HOMA2-IR and RC values for increased risk of cardiovascular mortality^24^. To assess the effect of LDL-c levels on our observation, we further stratified analyses by levels of LDL-c, comparing subjects above and below a threshold of 130mg/dL and by triglyceride levels, comparing subjects above and below a threshold of 200mg/dL.

#### IR as a mediator of the effect of remnant cholesterol on cardiovascular mortality

To assess the role of IR as mediator on the relationship between RC and cardiovascular mortality, we conducted causal mediation analyses with casually ordered mediators, using β-coefficients derived from Fine & Grey competing hazard regression models as proposed previously^25,26^. From mediation analyses, we estimated the average direct effect (ADE), the average causal mediation effects (ACME), the total effect and the proportion of the effect attributable to the mediator. To allow inference of the estimated parameters, we calculated 95% confidence intervals (95%CI) using accelerated non-parametric bootstrap percentiles with bias correction (B=1000). All statistical analyses were conducted using R software version 4.2.0.

## RESULTS

### Study population and mortality rates

From the initial sample of 13,867 subjects ≥20 years in NHANES-III and 17,570 in NHANES-IV, we included 16,201 subjects without diabetes with complete mortality follow-up, laboratory, and/or anthropometry data in whom it was possible to calculate RC (**Figure 1**). The analyzed sample was 52.3% men, had a median age of 45 (IQR 31-65) years, and a BMI of 26.2 (IQR 23.1-29.9) Kg/m^2^. **Table 1** shows the baseline characteristics of subjects included in the analysis, stratified by presence of IR. Compared with subjects without IR, individuals with IR who were alive at follow-up were older, were more likely to be male, had a higher number of comorbidities, and had a higher waist-to-height ratio (WHR), indicative of a higher degree of central obesity.

**Table 1.** Clinical parameters, all-cause and cause-specific mortality in subjects without diabetes, with and without insulin resistance (HOMA2-IR ≥2.5) included in the study.

| Parameter | Without IR (n=13,343) | With IR (n=2,858) | p-value |
| --- | --- | --- | --- |
| Age (years) | 41.0 (30.0-61.0) | 53.0 (37.0-69.0) | <0.001 |
| Male sex (%) | 6,248 (46.8) | 1,439 (50.3) | <0.001 |
| Follow-up time (Months) | 244.0 (155.0-293.0) | 192.0 (136.0-277.0) | <0.001 |
| Body-mass index (kg/m<sup>2</sup>) | 25.43 (22.60-28.80) | 30.50 (27.09-35.0) | <0.001 |
| Fasting glucose (mg/dL) | 90.0 (84.0-96.0) | 98.0 (91.0-106.0) | <0.001 |
| HbA1c (%) | 5.2 (5.0-5.5) | 5.5 (5.2-5.8) | <0.001 |
| Prediabetes (%) | 2227 (11.5) | 913 (26.2) | <0.001 |
| Smoking (%) | 3519 (26.4) | 674 (23.6) | <0.001 |
| Hypertension (%) | 2,892 (21.7) | 1,151 (40.3) | <0.001 |
| HDL-c (mg/dL) | 51.0 (43.0-62.0) | 43.0 (36.0-52.0) | <0.001 |
| LDL-c (mg/dL) | 124.58 (101.14) | 129.97 (105.82-155.82) | <0.001 |
| Remnant cholesterol (mg/dL) | 17.96 (13.29-25.67) | 28.29 (20.33-40.49) | <0.001 |
| Remnant cholesterol $\geq 30$mg/dL (%) | 2,343 (17.6) | 1,301 (45.5) | <0.001 |
| All-cause mortality (%) | 3,571 (26.8) | 1,181 (41.3) | <0.001 |
| Cardiovascular mortality (%) | 718 (5.4) | 284 (9.9) | <0.001 |
| Cerebrovascular mortality (%) | 213 (1.6) | 82 (2.9) | <0.001 |

Abbreviations:

### Mortality rates overall and according to remnant cholesterol and insulin resistance levels

We observed an overall rate of all-cause mortality of 16.3 deaths per 1,000 person-years (95%CI 15.9-16.8) over 3,493,300 person-years of follow-up (**Table 2**). When stratifying by the presence of IR, all-cause, cardiovascular, cerebrovascular and ASCVD mortality were higher in individuals with IR; furthermore, individuals with RC ≥30mg/dL had significantly higher rates of all-cause, cardiovascular and ASCVD mortality, and non-significantly higher rates of cerebrovascular mortality (**Table 2**, **Supplementary Material**). When combining categories of IR and RC levels, individuals with IR, independent of RC levels, had higher rates of all-cause, CVD, cerebrovascular and ASCVD mortality compared to individuals without IR and normal RC levels. Individuals without IR but with RC ≥30mg/dL had significantly higher rates of all-cause and ASCVD mortality compared to individuals without IR and RC<30mg/dL (**Figure 2**). These trends suggest that the impact of IR predominates over RC for ASCVD mortality in subjects without diabetes; nevertheless, elevated RC levels are still associated with higher rates of all-cause and ASCVD mortality.

**Figure 2.**
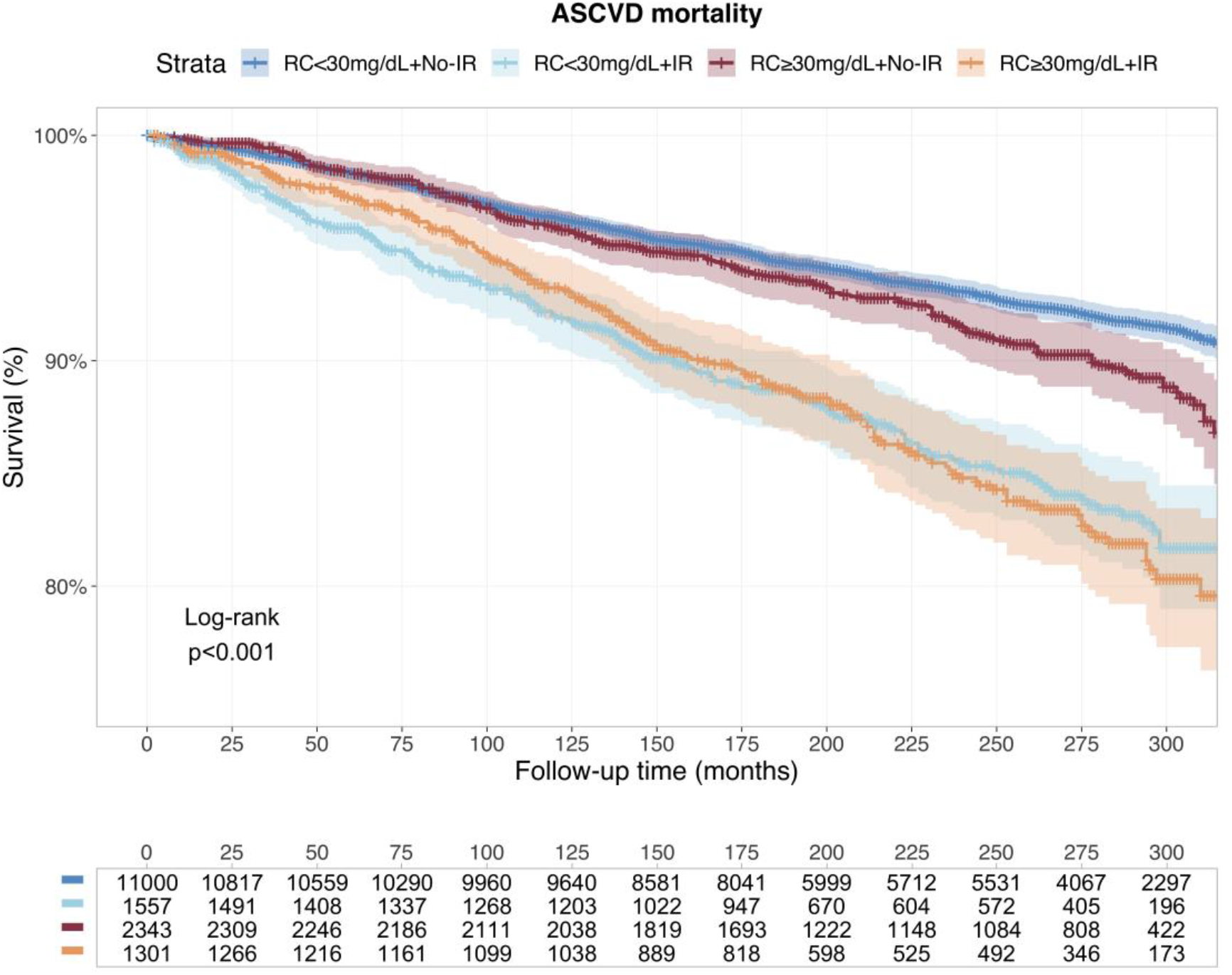
Kaplan-Meier curves for ASCVD mortality in individuals evaluated in NHANES-III and IV stratified according to joint remnant cholesterol levels (RC) ≥30mg/dL to define elevated remnant cholesterol and HOMA2-IR ≥2.5 to define insulin resistance. *<u>Abbreviations:</u>*RC: Remnant-Cholesterol calculated with Sampson’s LDL-C formula; HOMA2-IR: Homeostasis Model Assessment for Insulin Resistance; LDL-c: Low-density lipoprotein cholesterol.

**Table 2.** All-cause and cause-specific mortality rates (deaths per 1,000 person-years) in subjects without diabetes included in the study. <u>Abbreviations:</u> ASCVD, Atherosclerotic cardiovascular disease; IR, insulin resistance; RC, remnant cholesterol.

| Sample (n) | All-cause mortality<br>(95%CI) | Cardiovascular<br>mortality (95%CI) | Cerebrovascular<br>mortality (95%CI) | ASCVD mortality<br>(95%CI) |
| --- | --- | --- | --- | --- |
| Overall (n=16,201) | 16.32 (15.86-16.79) | 3.44 (3.23-3.66) | 1.01 (0.90-1.13) | 4.46 (4.21-4.70) |
| Without IR (n=13,343) | 14.58 (14.10-15.06) | 2.93 (2.72-3.15) | 0.86 (0.75-0.99) | 3.80 (3.56-4.05) |
| With IR (n= 2,858) | 25.54 (24.09-27.00) | 6.14 (5.43-6.86) | 1.77 (1.39-2.16) | 7.91 (7.11-8.73) |
| RC <30mg/dL (n=12,557) | 15.19 (14.68-15.70) | 3.17 (2.94-3.40) | 0.93 (0.81-1.06) | 4.10 (3.84-4.37) |
| RC ≥30mg/dL (n=3,644) | 30.38 (19.27-21.49) | 4.41 (3.90-4.93) | 1.30 (1.02-1.58) | 5.71 (5.13-6.30) |
| No-IR + RC <30mg/dL<br>(n=11,000) | 13.93 (13.42-14.44) | 2.81 (2.58-3.04) | 0.83 (0.71-0.96) | 3.64 (3.38-3.91) |
| No-IR + RC ≥30mg/dL<br>(n=1,557) | 17.72 (16.46-19.00) | 3.52 (2.96-4.09) | 1.04 (0.73-1.35) | 4.56 (3.92-5.21) |
| IR + RC <30mg/dL<br>(n=2,343) | 25.49 (23.50-27.48) | 6.12 (5.15-7.09) | 1.73 (1.21-2.25) | 7.85 (6.75-8.95) |
| IR + RC ≥30mg/dL<br>(n=1,301) | 25.61 (23.47-27.76) | 6.17 (5.12-7.22) | 1.82 (1.25-2.39) | 7.99 (6.79-9.19) |

### Relationship between remnant cholesterol and insulin resistance

We identified a significant association between higher HOMA2-IR and a higher RC concentration, independent of age, sex, BMI, ethnicity, HbA1c, number of comorbidities, smoking, HDL-C and LDL-C (**Figure 3A, Supplementary Material**). We also explored RC levels in subjects with and without IR and identified significantly higher RC values in subjects with IR (33.28±19.15 vs. 21.65±13.22, p<0.001); interestingly, higher RC persisted in IR after stratifying according to LDL-c and triglyceride values (**Figure 3B, Supplementary Material**). RC also predicted higher HOMA2-IR levels in fully adjusted models, which suggests that the relationship is bidirectional (**Supplementary Material**).

**Figure 3.**
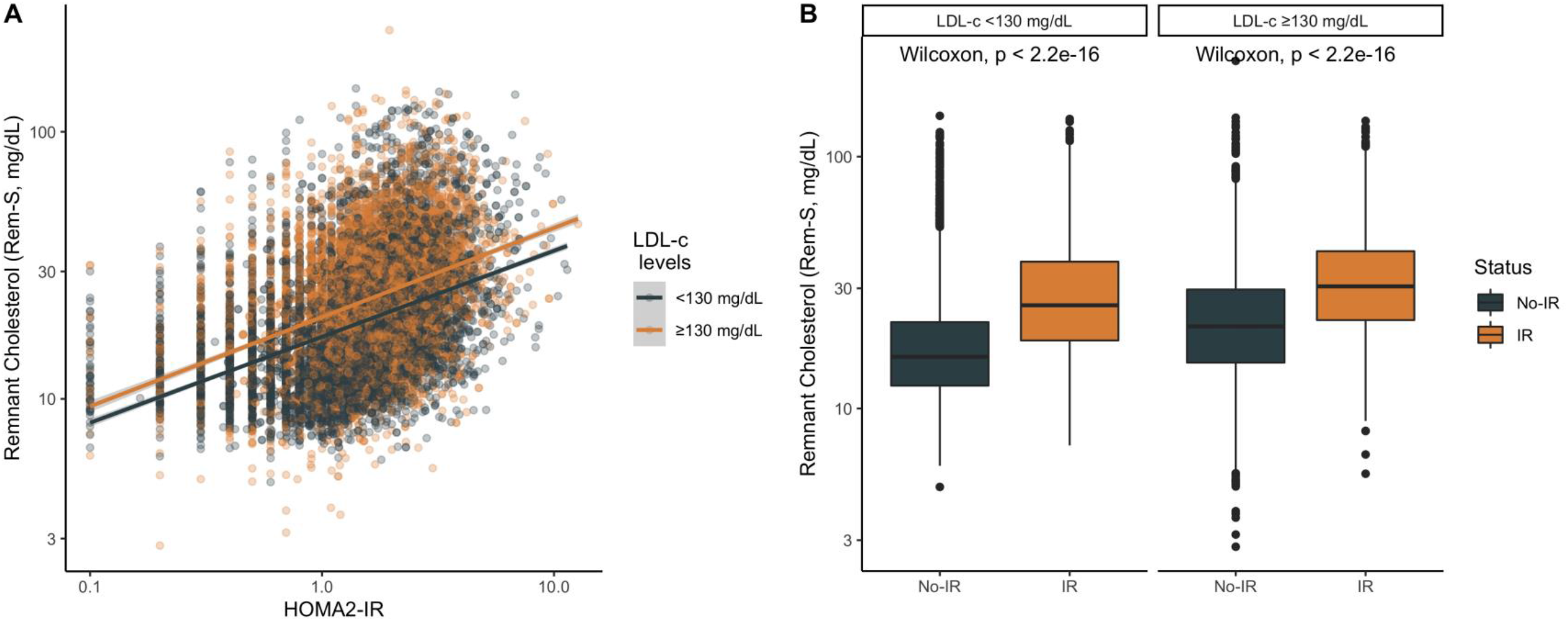
Relationship between remnant cholesterol calculated using Sampson’s formula for LDL-c (Rem-S) and HOMA2-IR in subjects without diabetes according to LDL-c levels, using values ≥130 mg/dL as a threshold to define high LDL-c (A). We also compared remnant cholesterol levels in subjects with and without insulin resistance (IR, HOMA2-IR ≥2.5) according to high vs. normal LDL-c levels (B). *<u>Abbreviations:</u>* Rem-S: Remnant-Cholesterol calculated with Sampson’s LDL-C formula; HOMA2-IR: Homeostasis Model Assessment for Insulin Resistance; ASCVD mortality, Atherosclerotic cardiovascular disease mortality.

### Association of remnant cholesterol and insulin resistance with cardiovascular mortality

We observed an increase in the risk of ASCVD mortality with increasing standard deviations of RC compared to other causes of death in univariate analyses (sHR 1.14, 95%CI 1.10-1.19, p<0.001, **Figure 4A**). When considering a model additionally comprising HDL-C and LDL-C, we identified a significant interaction between LDL-C and RC, whereby risk associated with RC decreased at higher levels of LDL-C (**Figure 4C**). Further adjustments by covariates observed a predominance of LDL-C over RC, without a significant interaction between RC and LDL-C but with an interaction between higher RC values and increasing age. Finally, when including HOMA2-IR into the model, RC and any interactions did not remain significant, whereas both HOMA2-IR and the interaction between HOMA2-IR and age remained significant (**Figure 4C**). HOMA2-IR values were also associated with higher risk of ASCVD mortality in both univariate (sHR 1.29, 95%CI 1.24-1.34, p<0.001, **Figure 4B**) and in models adjusted for sex, BMI, ethnicity, smoking, hypertension, LDL-c, HDL-C, HbA1c levels and number of comorbidities (sHR 1.68, 95%CI 1.42-1.98, p<0.001), with a similar interaction with increasing ages (sHR 0.993, 95%CI 0.991-0.995, p<0.001). These models were similar using RC calculated with LDL-C derived from Martin’s formula (**Supplementary Material**).

**Figure 4.**
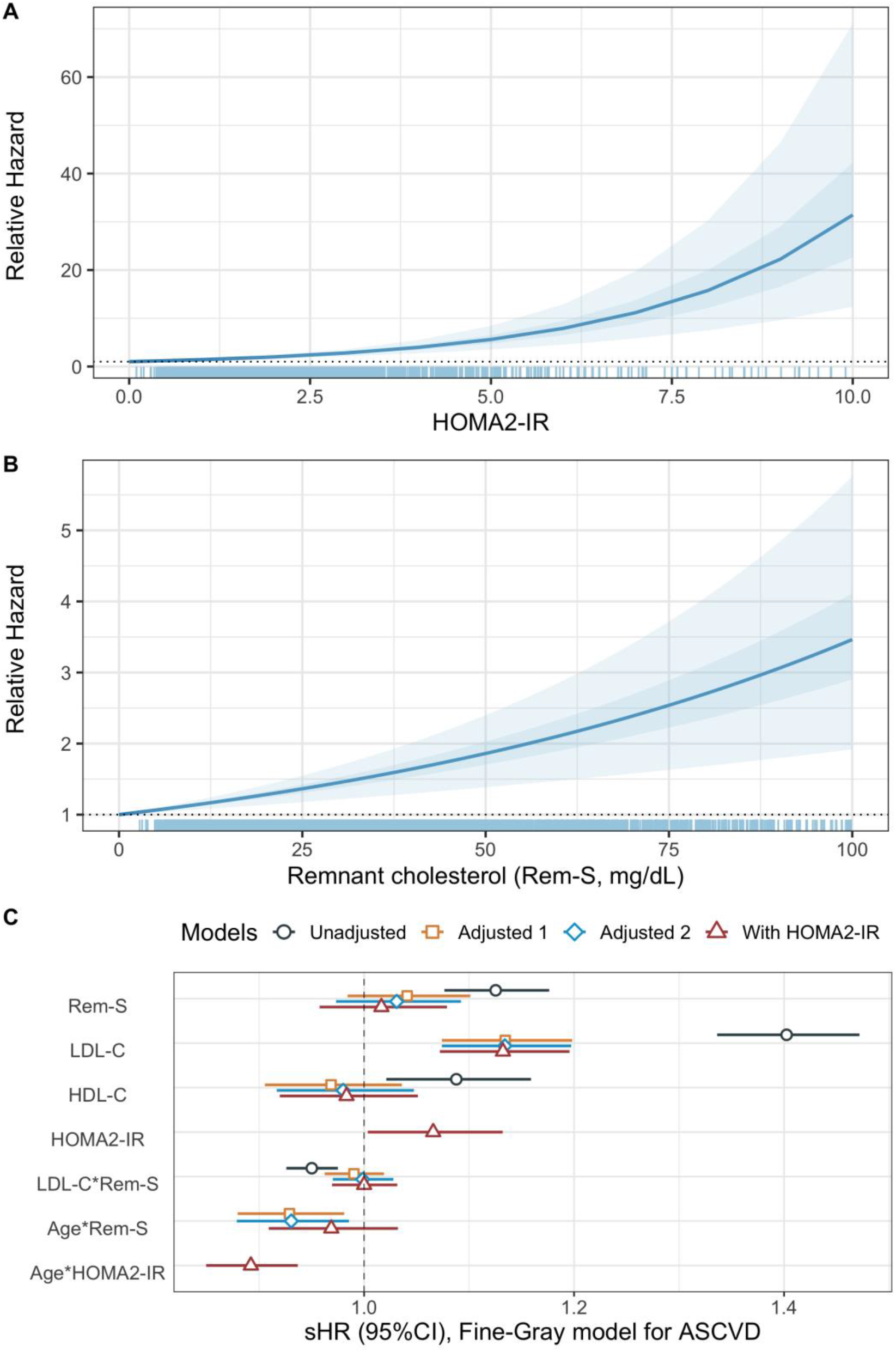
Relative hazard from Fine & Gray competing-risk regression model to assess the risk of ASCVD mortality driven by increased HOMA2-IR (A) and elevated remnant cholesterol levels (B) using the *simPH* R package. Sub distribution hazard ratios and corresponding 95%CI for ASCVD mortality (C) included sequentially with only lipoproteins, additionally adjusted for age and sex (Adjusted 1), for body-mass index, ethnicity, smoking, hypertension, LDL-c, HDL-C, HbA1c levels and number of comorbidities (Adjusted 2) and finally including HOMA2-IR. Interactions are shown with the (*) sign. *<u>Abbreviations:</u>* ASCVD, Atherosclerotic cardiovascular disease; Rem-S: Remnant-Cholesterol calculated with Sampson’s LDL-C formula; HOMA2-IR: Homeostasis Model Assessment for Insulin Resistance.

### Joint effect of remnant cholesterol and insulin resistance on cardiovascular mortality

When fitting a model with both RC and HOMA2-IR values together without adjustment, there was an interaction where the risk of cardiovascular mortality attributable to higher RC values decreased with increasing values of HOMA2-IR (sHR 0.934, 95%CI 0.897-0.973, p=0.001). Notably, in the fully adjusted model, significance for RC was not achieved (sHR 1.020, 95%CI 0.965-1.078, p=0.482), but higher risk persisted for higher HOMA2-IR values (HR 1.667, 95%CI 1.413-1.967, p<0.001), as well as an interaction where the risk of ASCVD mortality attributable to higher HOMA2-IR values decreased with increasing age (sHR 0.993, 95%CI 0.991-0.996, p<0.001, **Figure 4C**). When assessing combinations of high RC (≥30mg/dL) with IR (HOMA2-IR ≥2.5), we identified a higher risk of cardiovascular mortality in subjects with IR but normal RC (sHR 1.19, 95%CI 1.01-1.41, p=0.037) and in subjects with IR and high RC (sHR 1.27, 95%CI 1.07-1.50, p=0.006), but not in subjects without IR and high RC (sHR 0.965, 95%CI 0.826-1.126, p=0.648), compared to subjects without IR and RC in normal ranges in the fully adjusted model. This likely indicates that the effect of RC on ASCVD mortality was party mediated by a state of IR in individuals without diabetes, even when accounting for LDL-c values, which persist as risk factors for ASCVD mortality. In sensitivity analyses, models were conducted additionally using RC calculated from LDL-c estimated using Martin’s formula, yielding similar results (**Supplementary Material**).

### Insulin resistance mediates the effect of remnant cholesterol on cardiovascular mortality

To unravel the extent to which IR mediated the increased risk for ASCVD mortality observed with higher RC values, we fitted causal mediation models with causally ordered mediators to test the hypothesis that insulin resistance would mediate the effect of remnant cholesterol on cardiovascular mortality. We observed that the average direct effect of RC on cardiovascular mortality was not significant (ADE 1.001, 95%CI 0.998-1.005); however, the average mediating effect of IR (ACME 1.008, 95%CI 1.006-1.011, p<0.001) was associated with an increase in the relative incidence of cardiovascular mortality in subjects without diabetes. Therefore, IR accounts for 86.5% (95%CI 78.8-100.0) of the effect of increasing values of RC on cardiovascular mortality, likely indicating that IR is a complete mediator of the effect of RC on cardiovascular mortality. Most notably, when analyzing RC as a mediating variable instead of HOMA2-IR, only the direct effect of HOMA2-IR on ASCVD was significant, whilst the mediating pathway involving RC was not, suggesting that the impact of HOMA2-IR is independent of RC (**Supplementary Material**). As a sensitivity analysis, we refitted the model considering LDL-C as the effector variable instead of remnant cholesterol. Interestingly, the direct effect of LDL-C on ASCVD mortality was significant (ADE 1.003, 95%CI 1.002-1.004), but it was not mediated by IR (ACME 0.9999, 95%CI 0.9998-1.0001), indicating that the mediating mechanism of IR for the risk of ASCVD mortality is only relevant for RC.

**Figure 5.**
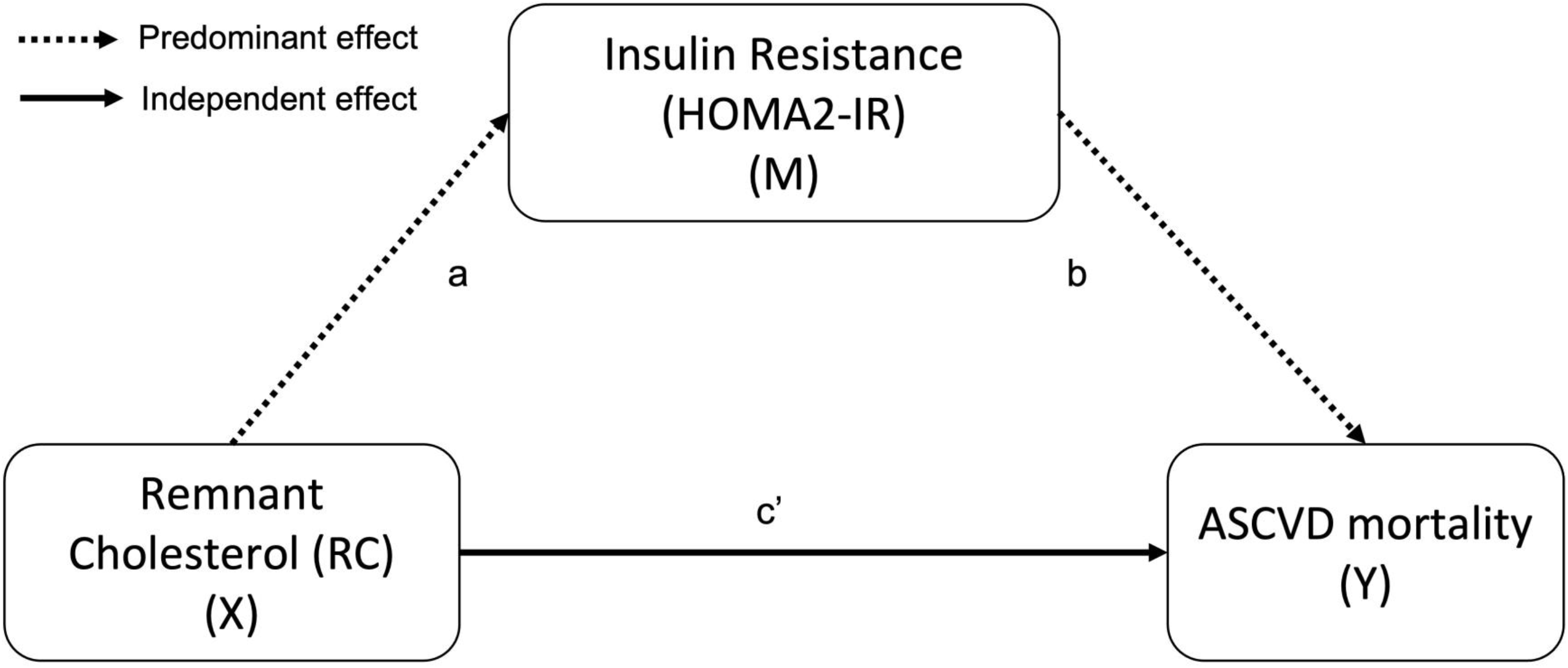
Proposed mechanism describing the relationship between remnant cholesterol (RC, effector X) and insulin resistance defined by HOMA2-IR levels (mediator M) in ASCVD mortality (outcome Y). Our results suggest that the effect of RC-induced insulin resistance on ASCVD mortality (pathway *a* − *b*) predominates over the direct effect of RC (pathway *c*’). *<u>Abbreviations:</u>* ASCVD, Atherosclerotic cardiovascular disease; RC: Remnant-Cholesterol calculated with Sampson’s LDL-C formula; HOMA2-IR: Homeostasis Model Assessment for Insulin Resistance.

## DISCUSSION

In this study we propose a mechanistic model that characterizes the relationship between IR, remnant cholesterol levels and ASCVD mortality in individuals without diabetes. Our results show that the effect of cholesterol remnants exerted on cardiovascular mortality is partly mediated by its effect in whole-body insulin sensitivity in subjects without diabetes; similarly, a state of IR may increase RC levels, thereby potentiating its associated cardiovascular risk. Furthermore, we show that all-cause, cardiovascular, cerebrovascular and ASCVD mortality increase in subjects with IR and in those with RC ≥30mg/dL; notably, this effect is primarily driven by a state of IR which is modified by age, whereby increasing age leads to decreases in the risk of ASCVD mortality conferred by IR. Our results suggest a need for further evidence from randomized control trials investigating whether reducing RC and improving insulin sensitivity would provide significant long-term cardiovascular benefits; moreover, given our observation that the joint effect of RC and IR on CVD risk seems to be more significant in younger adults these mechanisms should be further explored in this population.

Several studies have shown that the cholesterol content of TRL and their remnants are associated with cardiovascular risk and/or mortality, even in patients with optimal LDL-c levels regardless of apolipoprotein B-100 levels^6,21,27,28^. Furthermore, in patients with IR, cardiovascular risk is increased with exposure time, independent of LDL-C levels^29^. Our results show that both the cholesterol content in remnants and IR are associated with a higher risk of all-cause, cardiovascular, and cerebrovascular mortality in subjects without diabetes; however, a joint evaluation of both IR and RC shows a predominant effect of IR over RC in the risk of ASCVD, with a significant additional contribution of elevated LDL-C levels. These findings support the growing evidence suggesting that triglyceride and/or cholesterol content within TRLs, which are markedly increased in IR states, may contribute to the residual risk of ASCVD in addition to the atherogenic potential of RC^30–32^. Although the relationship between triglycerides and ASCVD is controversial, the presence of hypertriglyceridemia is associated with alterations in lipoprotein composition, low HDL-c and high non-HDL-c levels; which includes small, dense, and total LDL particles, intermediate density lipoprotein (IDL), and apolipoprotein C-III, all of which are associated with an increased risk of cardiovascular disease and mortality^3,31,33,34^. Based on our results, we propose that the presence of IR potentiates the atherogenic profile of RC, which are likely to exert a greater effect in younger individuals with lower LDL-C levels.

The relationship between RC and IR has clinical and pathophysiological significance in the development and clinical progression of cardiovascular disease. Previous studies have shown that the development of atherosclerotic lesions in patients at high cardiovascular risk are impacted by both RC and IR, even when taking into consideration glycemia^10,35^. Furthermore, studies have shown that the VLDL receptor, which also functions as a RC receptor, has a key role in obesity, IR, and hyperlipidemia in mice fed a high-fat, refined sugar diet (HFS). Interestingly, in VLDL receptor knockout mice that receive HSF, there is an increase in plasma VLDL remnants; however, VLDLR knockout mice are not capable of storing triglycerides in adipose tissue and, despite HSF, they do not develop IR. Therefore, the formation of VLDL remnants due to HSF could be considered a precursor to obesity and IR^36–38^. In humans, previous studies have shown that mild acute hypertriglyceridemia leads to impaired oral glucose tolerance and reduced whole-body insulin sensitivity, which suggest a potential role of TRL and its remnants in inducing a state of IR^39^. This mechanism may explain the mediating role of IR in increasing RC-mediated cardiovascular risk observed in our study, considering that the mechanism whereby increased RC levels lead to ASCVD mortality may be through the direct effect of RC on insulin action, with subsequent joint effects in increasing risk of ASCVD mortality.

### Study limitations

Our work has some strengths and limitations. First, by using a large and ethnically diverse population-based study we were able to investigate the joint impact of RC and IR on ASCVD mortality in individuals without diabetes. Despite this strength, due to the cross-sectional nature of NHANES, it was not possible to retrieve information on clinical, biochemical and/or anthropometric characteristics during follow-up; therefore, the risk estimate is based on a single baseline measurement, which may not reflect the dynamic interplay between insulin sensitivity, lipoprotein metabolism and cardiovascular health. Despite the lack of a direct measurement of RC to establish a gold standard, RC levels were calculated using two formulas in our study -Sampson’s and Martin’s -both of which have adequate performance for LDL-c estimation and thus would lead to better estimates of RC in the setting of hypertriglyceridemia^19,20^. Furthermore, it was not possible to include additional variables that impact RC levels, such as treatment with fibrates and/or statins, which could modify the observed effect of RC in our study. We should also note that HOMA2-IR is a proxy of whole-body insulin resistance and does not indicate tissue-specific IR, which may have differential effect on both lipoprotein metabolism and cardiovascular health. Furthermore, given that the performance of HOMA2-IR to capture IR may be reduced in individuals with diabetes, particularly those on insulin therapy, further studies are required to investigate this phenomenon in individuals with diabetes and in those with extreme forms of hypertriglyceridemia, settings which are likely to modify the proposed pathophysiological pathways and thus differentially impact cardio-metabolic risk associated with RC and IR.

### Conclusions

We propose that IR partly mediates the effect of RC on cardiovascular disease and ASCVD mortality in individuals without diabetes. Our findings show that the effect of elevated RC contributes to increased risk of ASCVD mortality likely through both a direct effect on vascular health, and an indirect effect through the impact of elevated RC on IR. Notably, we also identified that in individuals without whole-body IR the impact of RC on ASCVD mortality is modest and atherogenicity is likely driven by elevated LDL-C levels. Most notably, the effect of IR on cardiovascular risk is modified by age, whereby its impact is stronger in younger individuals without diabetes. Given the significant impact attributable to RC in residual cardiovascular risk, future studies should investigate whether concomitant therapies to improve insulin sensitivity may provide an additional benefit in reducing cardiovascular disease burden and mortality.

## Data Availability

All code, datasets and materials are available for reproducibility of results at https://github.com/oyaxbell/remnant_cholesterol/

## ACKNOWLEDGMENTS

AVV and CAFM are enrolled at the PECEM Program of the Faculty of Medicine at UNAM. CAFM is supported by CONACyT. The graphical abstract and Figure 1 were designed using resources created by Flat Icon, Smashicons, Freepik, and Surang from www.flaticon.com

## AUTHOR CONTRIBUTIONS

Research idea and study design: AVV, CAFM, NEAV, LFC, DRG, GDL, JPDS, OYBC, JAS; data acquisition: AVV, OYBC, NEAV, CAFM, LFC; analysis/interpretation: AVV, OYBC, CAFM, NEAV, LFC; statistical analysis: AVV, OYBC; manuscript drafting: AVV, OYBC, CAFM, NEAV, LFC, DRG, GDL, JPDS, JAS, CAAS; supervision or mentorship: OYBC. Each author contributed important intellectual content during manuscript drafting or revision and accepts accountability for the overall work by ensuring that questions pertaining to the accuracy or integrity of any portion of the work are appropriately investigated and resolved.

