## Supplementary Material for "Insulin resistance potentiates the effect of remnant cholesterol on cardiovascular mortality in individuals without diabetes"

**SUPPORTING INFORMATION - Insulin resistance mediates the effect of triglyceride-rich lipoprotein remnants on cardiovascular mortality in individuals without diabetes.**

Arsenio Vargas-Vázquez, Carlos A. Fermín-Martínez, Neftali Eduardo Antonio-Villa, Luisa Fernández-Chirino, Daniel Ramírez-García, Gael Dávila-López, Juan Pablo Díaz-Sánchez, Carlos A. Aguilar-Salinas, Jacqueline A. Sieglie, Omar Yaxmehen Bello-Chavolla

**SUPPLEMENTARY FIGURES**

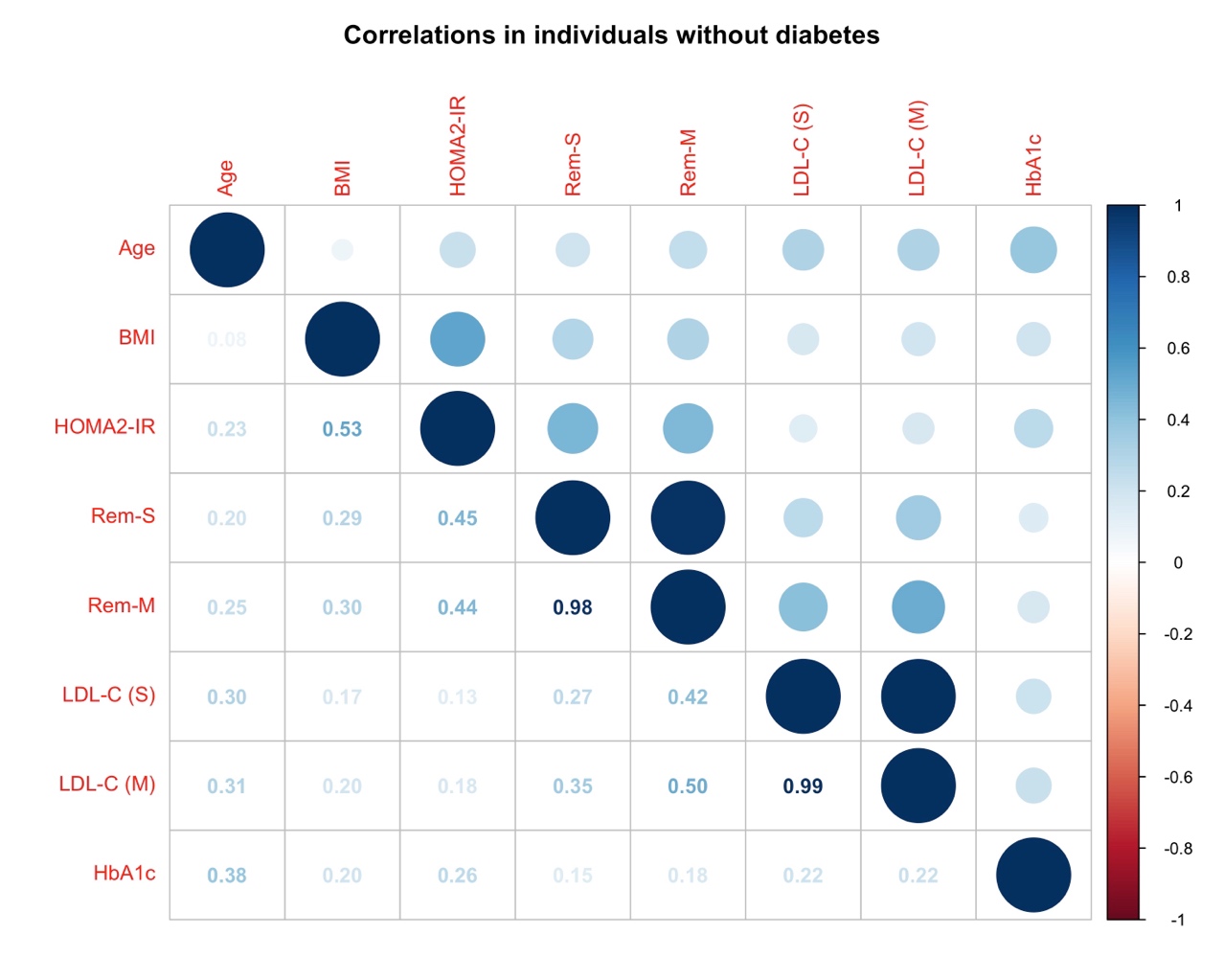

**Supplementary Figure 1.** Linear correlations between evaluated measurements calculated in NHANES-III and IV. Abbreviations: Rem-S: Remnant-Cholesterol calculated with Sampson’s LDL-C formula; Rem-M: Remnant-Cholesterol calculated with Sampson’s LDL-C formula; BMI: Body-mass index; LDL-C: Low-density lipoprotein cholesterol; HOMA2-IR: Homeostasis Model Assessment for Insulin Resistance.

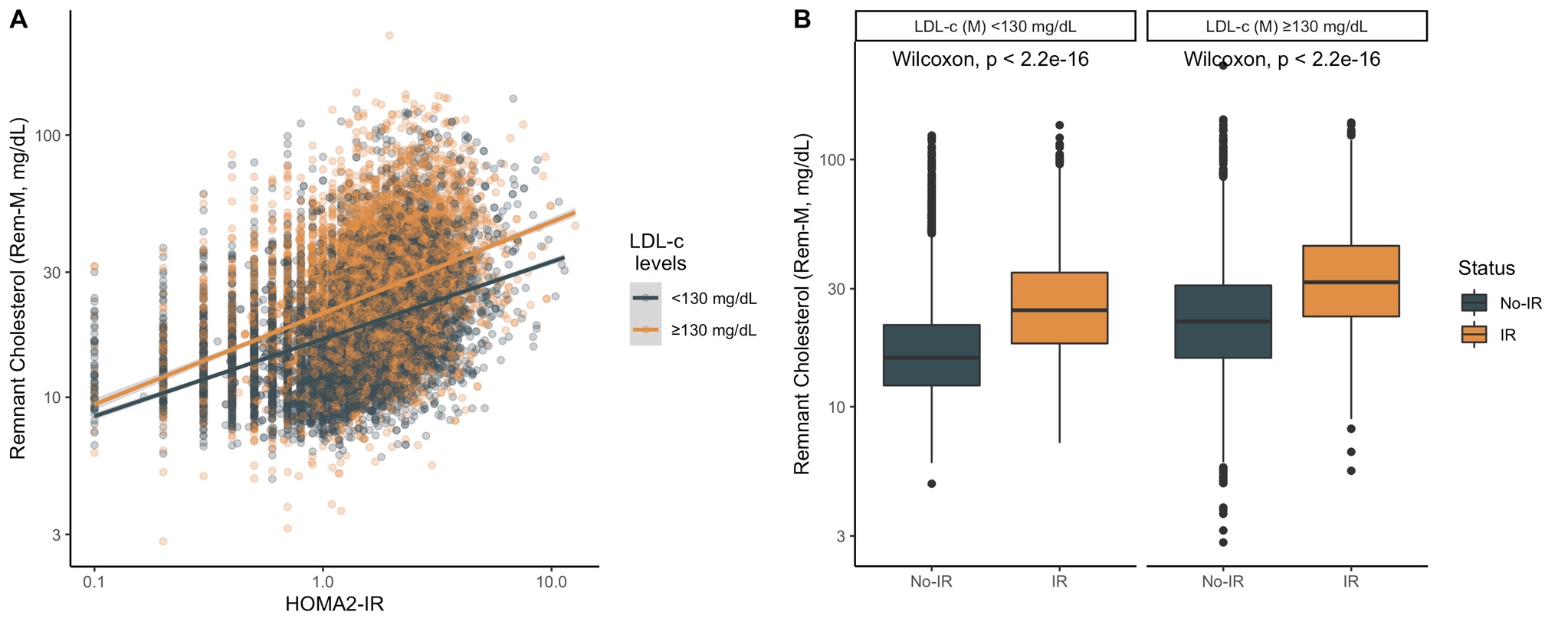

**Supplementary Figure 2.** Scatter plots of HOMA2-IR vs Remnant Cholesterol calculated using Martin’s formula for LDL (Rem-M, **A**). We also compared remnant cholesterol levels in subjects with and without insulin resistance (IR, HOMA2-IR ≥2.5) according to high vs. normal LDL-c levels (**B**). Abbreviations: Rem-M: Remnant-Cholesterol calculated with Martin’s LDL-C formula; HOMA2-IR: Homeostasis Model Assessment for Insulin Resistance; LDL-c: Low-density lipoprotein cholesterol.

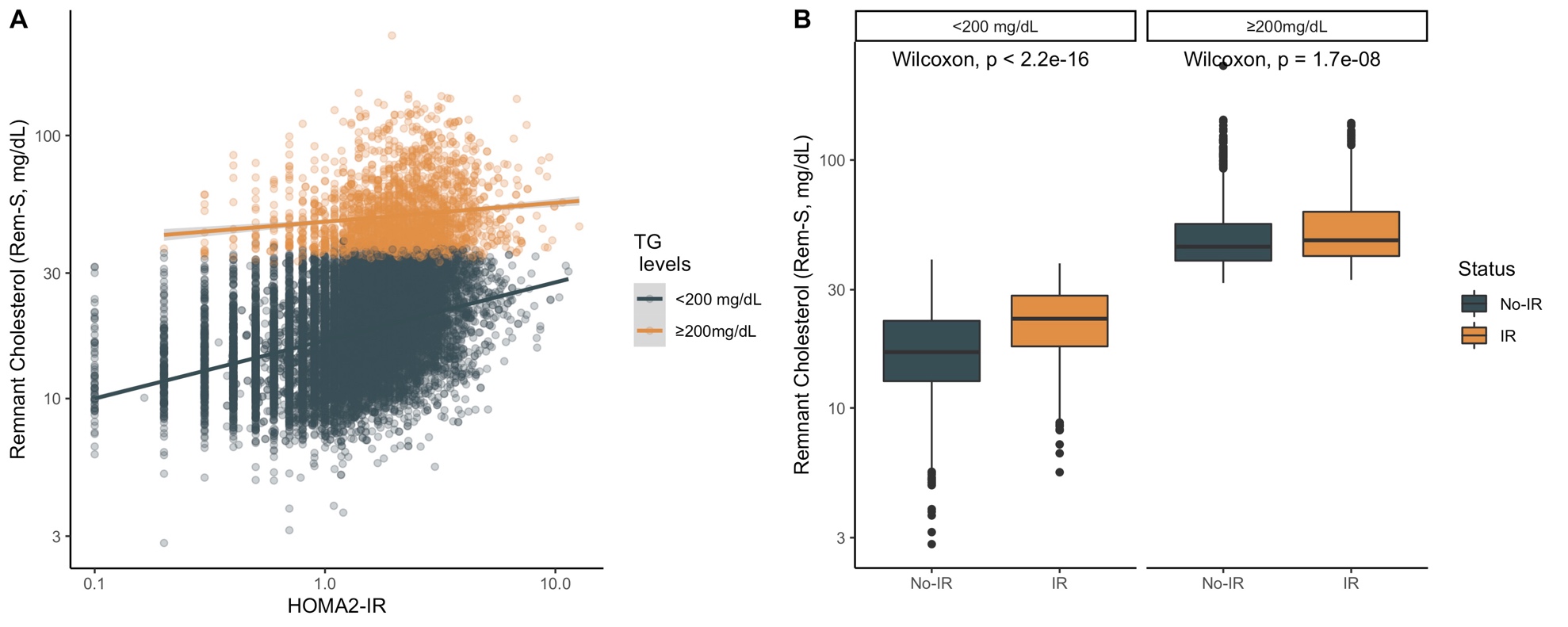

**Supplementary Figure 3.** Scatter plots of HOMA2-IR vs Remnant Cholesterol calculated using Sampson’s formula for LDL (Rem-M, **A**). We also compared remnant cholesterol levels in subjects with and without insulin resistance (IR, HOMA2-IR ≥2.5) according to high vs. normal triglyceride levels (**B**). Abbreviations: Rem-M: Remnant-Cholesterol calculated with Martin’s LDL-C formula; HOMA2-IR: Homeostasis Model Assessment for Insulin Resistance; TG: Triglycerides.

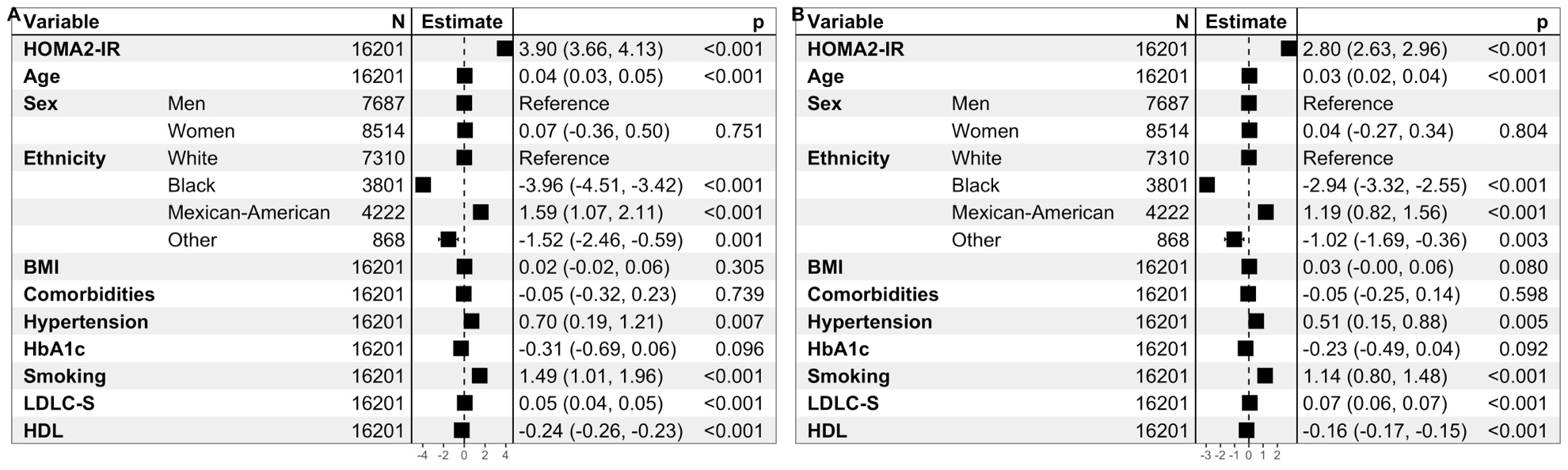

**Supplementary Figure 4.** Forrest plots of linear regression models for prediction of Remnant Cholesterol calculated using Sampson’s formula for LDL (Rem-S, **A**) and Martin’s formula for LDL-C (Rem-M, **B**) using HOMA2-IR and adjusted for covariates. Abbreviations: Rem-S: Remnant-Cholesterol calculated with Sampson’s LDL-C formula; Rem-M: Remnant-Cholesterol calculated with Sampson’s LDL-C formula; BMI: Body-mass index; LDL-C: Low-density lipoprotein cholesterol using either Sampson’s (S) or Martin’s formula (M); HOMA2-IR: Homeostasis Model Assessment for Insulin Resistance.

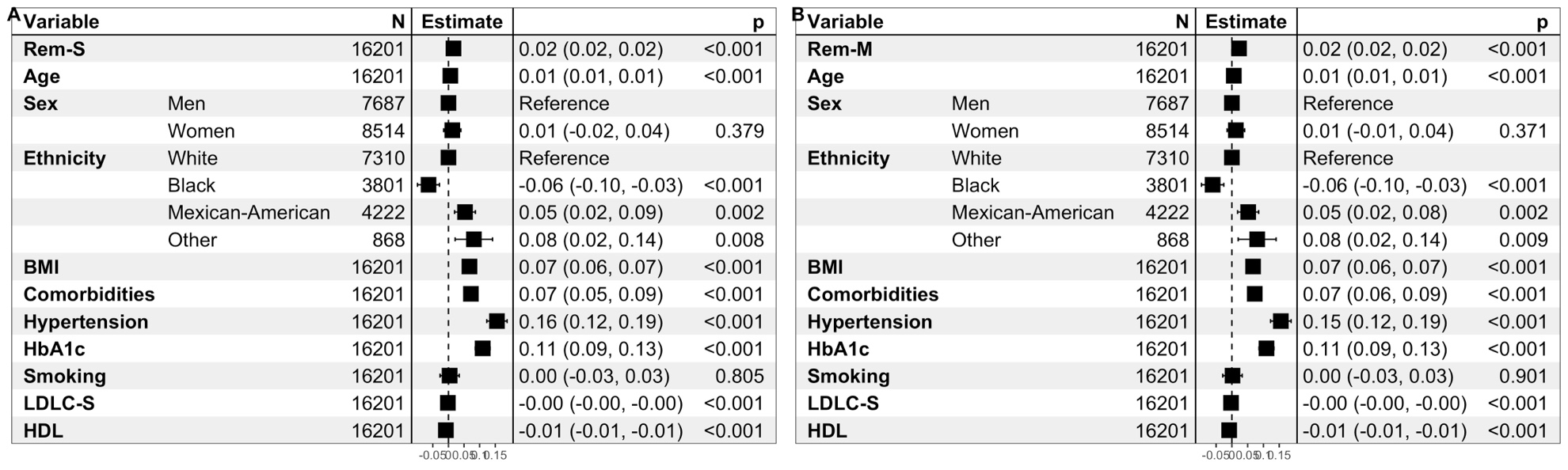

**Supplementary Figure 5.** Forrest plots of linear regression models for prediction of HOMA2-IR with Remnant Cholesterol calculated using Sampson’s formula for LDL (Rem-S, **A**) and Martin’s formula for LDL-C (Rem-M, **B**) adjusted for covariates. Abbreviations: Rem-S: Remnant-Cholesterol calculated with Sampson’s LDL-C formula; Rem-M: Remnant-Cholesterol calculated with Sampson’s LDL-C formula; BMI: Body-mass index; LDL-C: Low-density lipoprotein cholesterol using either Sampson’s (S) or Martin’s formula (M); HOMA2-IR: Homeostasis Model Assessment for Insulin Resistance.

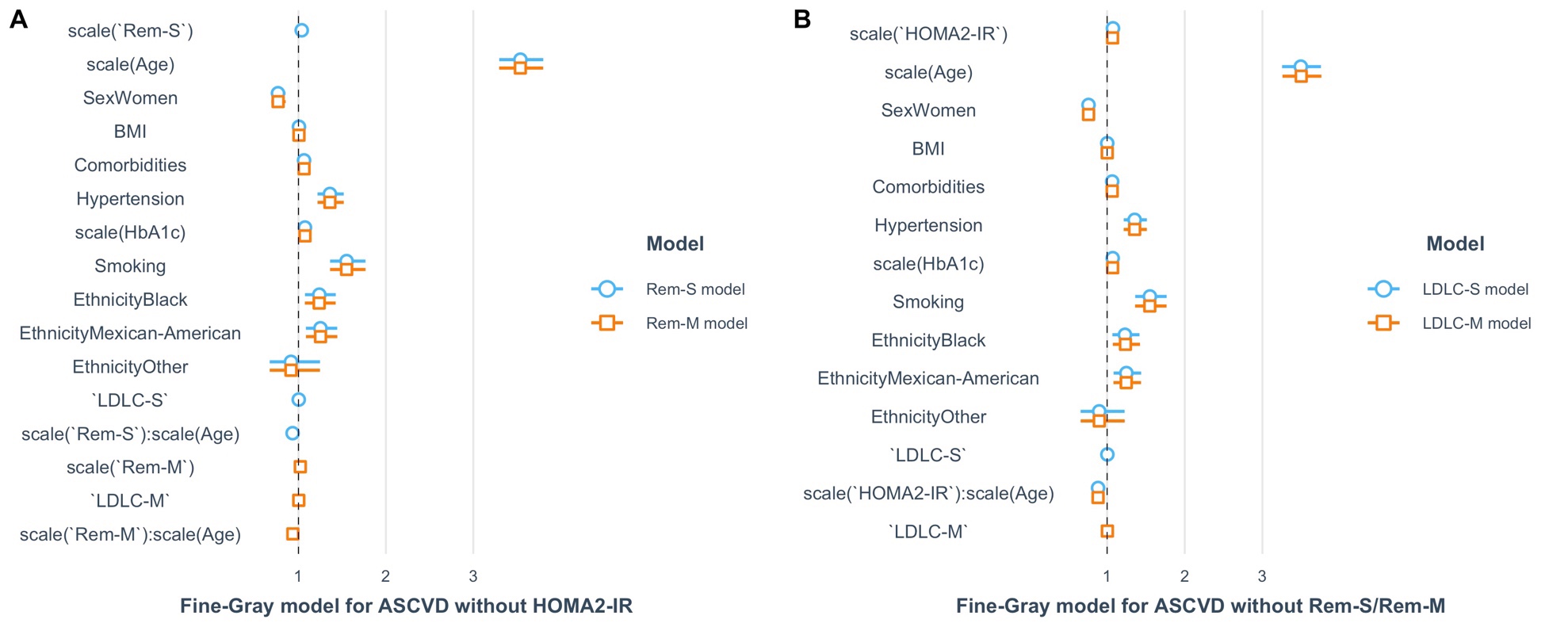

**Supplementary Figure 6.** Forrest plots of Fine & Gray competing-risk regression models for prediction of ASCVD using Remnant Cholesterol without HOMA2-IR (**A**) and without remnant cholesterol (**B**) comparing models using Rem-S/Rem-M and LDL-c calculated using Sampson’s (LDLC-S) and Martin’s (LDLC-M) LDL-C equations. Abbreviations: Rem-S: Remnant-Cholesterol calculated with Sampson’s LDL-C formula; Rem-M: Remnant-Cholesterol calculated with Sampson’s LDL-C formula; BMI: Body-mass index; LDL-C: Low-density lipoprotein cholesterol using either Sampson’s (S) or Martin’s formula (M); HOMA2-IR: Homeostasis Model Assessment for Insulin Resistance; ASCVD, Atherosclerotic cardiovascular disease. Results are presented as sub distribution Hazard Ratios (sHR).

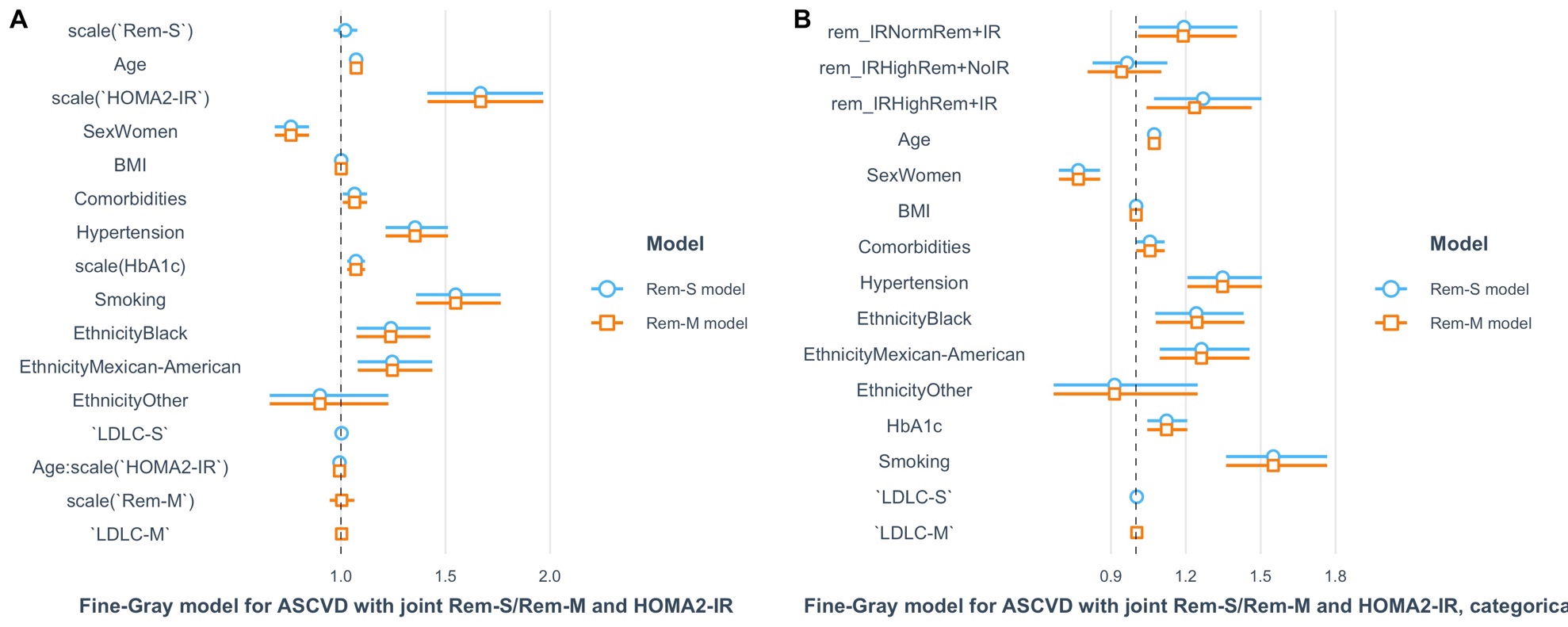

**Supplementary Figure 7.** Forrest plots of Fine & Gray competing-risk regression models for prediction of ASCVD using Remnant Cholesterol with HOMA2-IR (**A**) and using combinations of HOMA2-IR≥2.5 and Rem-S/Rem-M ≥30mg/dL (**B**) comparing models using Rem-S/Rem-M and LDL-c calculated using Sampson’s (LDLC-S) and Martin’s (LDLC-M) LDL-C equations. Abbreviations: Rem-S: Remnant-Cholesterol calculated with Sampson’s LDL-C formula; Rem-M: Remnant-Cholesterol calculated with Sampson’s LDL-C formula; BMI: Body-mass index; LDL-C: Low-density lipoprotein cholesterol using either Sampson’s (S) or Martin’s formula (M); HOMA2-IR: Homeostasis Model Assessment for Insulin Resistance; ASCVD, Atherosclerotic cardiovascular disease.

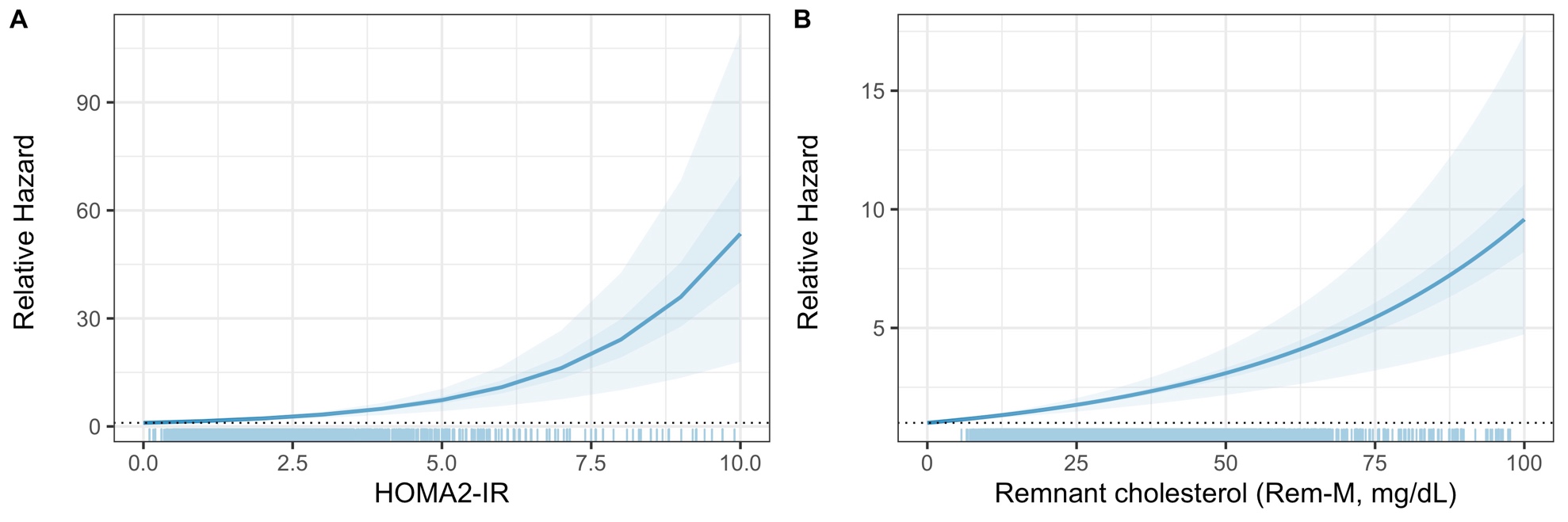

**Supplementary Figure 8.** Relative hazard from Fine & Gray competing-risk regression model to assess the risk of ASCVD mortality driven by increased HOMA2-IR and elevated remnant cholesterol levels estimated using Martin’s formula for LDL-c (Rem-M) using the simPH R package. Abbreviations: Rem-M: Remnant-Cholesterol calculated with Sampson’s LDL-C formula; HOMA2-IR: Homeostasis Model Assessment for Insulin Resistance; ASCVD, Atherosclerotic cardiovascular disease.

**SUPPLEMENTARY TABLE**

| Model | ADE (95%CI) | ACME (95%CI) | Total Effect (95%CI) | % Mediation (95%CI) |
| --- | --- | --- | --- | --- |
| Rem-S model | 1.001 (0.998-1.005) | 1.008 (1.006-1.011) | 1.010 (1.005-1.014) | 86.53 (78.76-100.0) |
| Rem-M model | 1.001 (0.995-1.006) | 1.012 (1.008-1.016) | 1.013 (1.006-1.019) | 97.35 (85.17-100.0) |
| Categorical Rem-S ≥30mg/dL model | 0.995 (0.882-1.123) | 4.052 (2.145-7.769) | 4.033 (2.119-7.787) | 100.0 (99.8-100.0) |
| Categorical Rem-M ≥30mg/dL model | 0.972 (0.860-1.100) | 4.050 (2.133-7.806) | 3.938 (2.057-7.649) | 100.0 (100.0-100.0) |

**Supplementary Table 1.** Causally ordered mediation models to evaluate the mediating role of HOMA2-IR (Rem-S/Rem-M model) and IR (HOMA2-IR ≥2.5, categorical models) in the relationship between remnant cholesterol and ASCVD. Abbreviations: Rem-M: Remnant-Cholesterol calculated with Sampson’s LDL-C formula; HOMA2-IR: Homeostasis Model Assessment for Insulin Resistance; ASCVD, Atherosclerotic cardiovascular disease.
